# Forecasting COVID-19, Influenza and RSV hospitalisations over winter 2023/24 in England

**DOI:** 10.1101/2024.09.07.24313175

**Authors:** Jonathon Mellor, Maria L. Tang, Owen Jones, Infectious Disease Modelling Team, Thomas Ward, Steven Riley, Sarah R. Deeny

## Abstract

Seasonal respiratory viruses cause substantial pressure on healthcare systems, particularly over winter. System managers can mitigate the impact on patient care when they anticipate hospital admissions due to these viruses. Hospitalisation forecasts were used widely during the SARS-CoV-2 pandemic. Now, resurgent seasonal respiratory pathogens add complexity to system planning. We describe how a suite of forecasts for respiratory pathogens, embedded in national and regional decision-making structures, were used to mitigate the impact on hospital systems and patient care.

We developed forecasting models predicting hospital admissions and bed occupancy two weeks ahead for COVID-19, influenza, and RSV in England over winter 2023/24. Bed occupancy forecasts were informed by the ensemble admissions models. Forecasts were delivered in real-time at multiple scales. The use of sample-based forecasting allowed for effective reconciliation and trend interpretation.

Admission forecasts, particularly RSV and influenza, showed high skill at regional levels. Bed occupancy forecasts had well-calibrated coverage, owing to informative admissions forecasts and slower moving trends. National admissions forecasts had mean absolute percentage errors of 27.3%, 30.9% and 15.7% for COVID-19, influenza, and RSV respectively, with corresponding 90% coverages of 0.439, 0.807 and 0.779.

These real-time winter infectious disease forecasts produced by the UK Health Security Agency for healthcare system managers played an informative role in mitigating seasonal pressures. The models were delivered regularly and shared widely across the system to key users. This was achieved by producing reliable, fast, and epidemiologically informed ensembles of models. Though, a higher diversity of model approaches could have improved forecast accuracy.

## 1. Introduction

In England, each year respiratory pathogens circulating in the community cause strain on the healthcare system, particularly SARS-CoV-2, influenza, and Respiratory Syncytial Virus (RSV). In the 2023/24 winter season there were 96.6, 76.9 and 37.8 cumulative admissions per 100,000 people in England with COVID-19, influenza, and RSV respectively [1] [2]. Public health and healthcare leaders can better use scarce resources when they can predict hospital admissions and bed occupancy, reallocating staff and beds to elective care as emergency admissions decline.

Forecasting is widely used to give quantitative assessments on future events, harnessing historic trends and modelled assumptions. However, infectious disease dynamics are challenging to confidently understand in real-time due to their complexity and partial observation. When used well, numerical forecasts can help inform healthcare planning to allocate system resources more effectively, leading to improved health outcomes. Within England the UK Health Security Agency (UKHSA) is responsible for the prevention, preparation, and response to infectious diseases. The National Health Service England (NHSE) provides healthcare provision to the population, and both government organisations are overseen by the Department of Health and Social Care.

Infectious disease forecasts have been produced for influenza over many years in the United States by the Centre for Disease Control (CDC) [3] [4]. Forecasting is not limited to influenza, with challenges covering West Nile Virus [5], Ebola [6], and Chikungunya [7], with collaborations between government and academia driving improvements in the field [8] [9]. These methods and initiatives were stepped up during the SARS-CoV-2 pandemic responding to increased demand [10] [11], used in communications by the US CDC to the public.

The SARS-CoV-2 pandemic disrupted seasonal patterns of many respiratory viruses, driven by changing population mixing dynamics caused by control interventions [12], notably limiting influenza and RSV transmission. As population mixing returned to pre-pandemic levels these diseases re-emerged in winter 2022/23 [13]. Rising emergency hospital admissions due to influenza, COVID-19, and RSV place direct resource pressure on hospitals [14]. These admissions take up in-patient beds, and the increased demand on hospitals reduces the planned and non-emergency care available. Therefore, the accurate prediction of infectious disease pressures helps to manage the wider healthcare system.

A range of metrics are commonly used to measure and understand seasonal respiratory disease waves, each tackling different policy questions: peak magnitude, peak timing, cumulative incidence, and incidence over time [4]. Comparisons between rapid collections and retrospective records give us confidence in our choice of metrics indicating suitability for real-time modelling [15]. In addition, we focus on test-positive diagnosis rather than syndrome to ensure modelling is specific to a pathogen, allowing us to model diseases separately [16].

Over the winter 2023/24 season UKHSA delivered a suite of forecasting models to estimate hospital admissions and bed occupancy trends of COVID-19, influenza, and RSV waves. This breadth of pathogens and metrics modelled was key to the suite’s utility. The forecasts were used directly to inform national policy discussions and integrated into regional level decision making. In this work we outline our approach in delivering this forecasting suite, key considerations, and describe our real-time results.

## 2. Methods

### 2.1 Data sources

The National Health Service (NHS) is a publicly funded healthcare system covering England, with data collection consistent across hospitals [17]. NHS England geographic structures are given in **Supplementary** Figure 1. Individuals presenting severe symptoms of an infectious disease are often tested in secondary care settings, with diagnostic tests reported to UKHSA via the Second-Generation Surveillance System (SGSS). There is a high ascertainment and coverage of SARS-CoV-2, influenza, and RSV testing in hospitals for differential diagnosis. However, this ascertainment can vary with demographics such as age [18] and across primary care, secondary care, and community settings [19]. More broadly, all-cause hospital admissions can be predicted by seasonal patterns and environmental effects [20]. However, presentations due to infectious disease are less predictable, particularly when seasonal transmission patterns are disrupted. Test-positive hospitalisations; admissions and bed occupancy, are key metrics for the pressures sustained by hospitals due to infectious disease. Forecast target definitions are provided (**Table 1**), highlighting their differences.

**Table 1.**
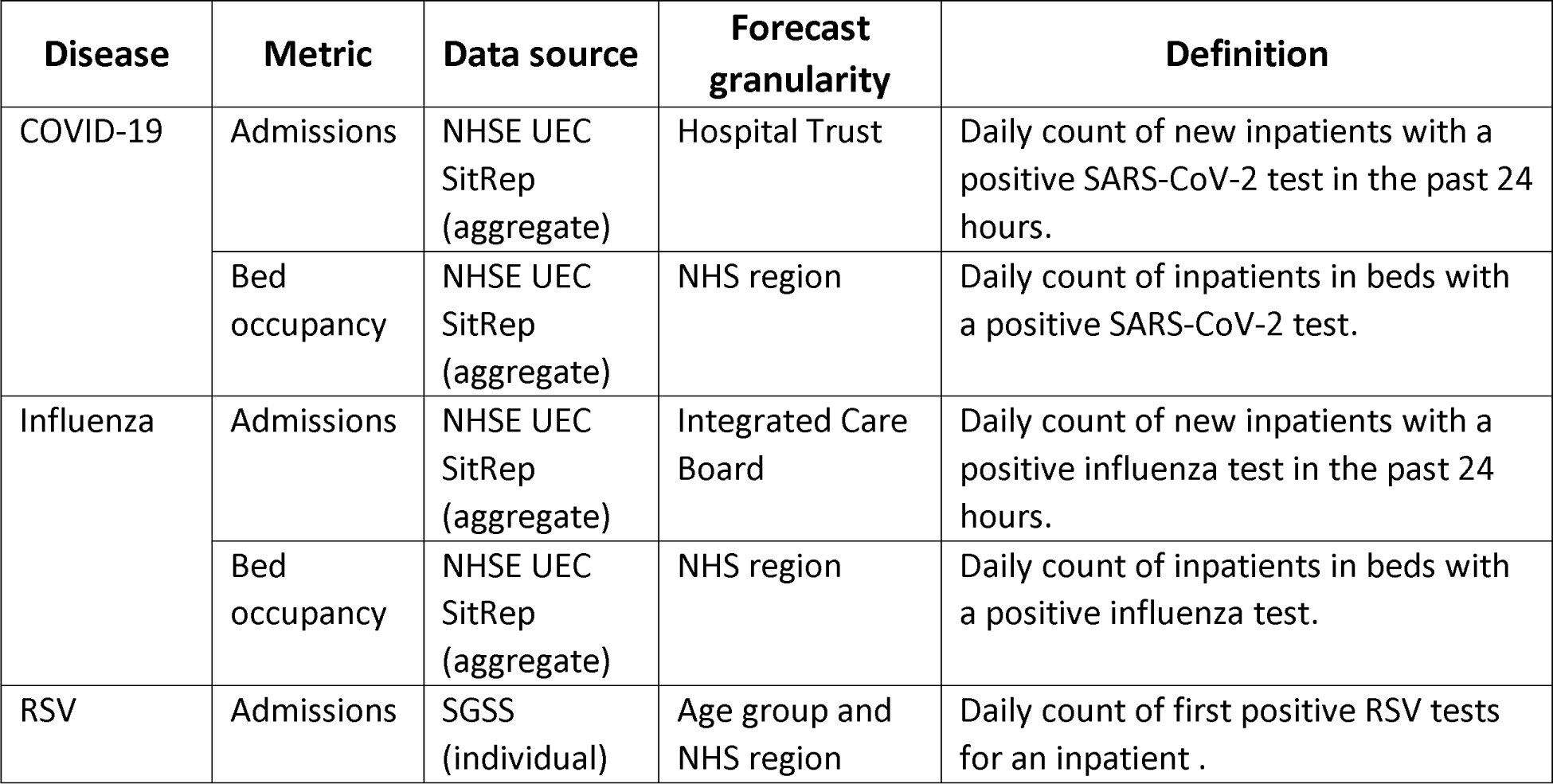
A breakdown of each forecasting target chosen across diseases and metrics. The National Health Service England (NHSE) Urgent and Emergency Care (UEC) Situational Report is a daily census of aggregate counts. The Second-Generation Surveillance Service (SGSS) is a national database for infectious disease diagnostic test results. Within the health geographies of England, a hospital Trust is a collection of hospitals nested within an Integrated Care Board (ICB), nested within an NHS region of the country. The geographic structures are further detailed in **Supplementary** Figure 1.

### 2.2 Forecast Models

#### Statistical and ensemble rationale

Our modelling suite focusses on statistical forecasts, aligning with the evidence of their strong performance in short-term forecasting tasks, relative to strictly mechanistic approaches [21] [22]. Alongside their predictive performance, this class of models has fast runtimes, are largely disease agnostic, and do not require substantial historic data. Furthermore, we developed model ensembles, which capture uncertainty in possible trends [23], with evidence of improved performance [24] [25]. Model ensembles also reduce our reliance on individual models, improving operational resilience.

#### Ensemble inclusion criteria

In a *M*-open model combination, for possible candidate models *M*, we ideally have a breadth of modelling approaches to produce combination estimates [26]. However, practical considerations are critical in ensemble design. Across pathogens and metrics, there is a single-day turnaround from modelling to dissemination, highlighting the importance of speed and reliability. Candidate models are selected if the model:

1. has a total runtime (fitting, inference, post-processing) across all locations for a disease in fewer than 15 minutes.
2. produces prediction samples (such as posterior samples).
3. improves forecasting capability.

Criteria 1 makes the production of all forecasts feasible, allowing for error flagging, model reruns, and thorough assurance of results. While models can be run in parallel, there are limitations in how many individuals are available to run the process, make fixes, and quality check results. Criteria 2 ensures desirable qualities for ensembling and coherence of prediction aggregations, such as from local to national levels. Criteria 3 is a holistic evaluation of model performance and utility. We do not select models based on scoring rules alone, but instead consider the wider properties of a model (reliability, ease-of-use, difference from existing models) and discussion with surveillance experts.

#### Specific model classes

A variety of statistical models were used to forecast trends, each with different underlying assumptions. Each model is structured separately for each disease, adapted to best reflect the surveillance data being modelled, with key hyperparameters retuned regularly over the season. The primary model class used across the pathogens was a Generalised Additive Model (GAM), which uses the semi-mechanistic assumption of a recent growth-rate extrapolated forward in time [27] via the *mgcv* and *gratia* packages [28] [29]. This model has different variations across the diseases, using different hierarchical components, geographies, and for the RSV model a structure pooling trends across adjacent age groups. To improve model performance, we leverage probabilistic catchment areas to define a denominator population for hospital admissions [30]. Secondly, we use state-space based models, primarily with an Error Trend Seasonality (ETS) structure fit to each individual geographic location’s time series via the *fable* package [31]. Lastly, syndromic surveillance is fed into regression models to predict expected future admissions given current leading indicators levels [32]. Models ran each week are first inspected by the modelling team (applying expert judgement), then candidate models are ensembled. Each model developed is given by disease in **Supplementary Table 1**.

For the COVID-19 and influenza admissions targets we developed a secondary forecast target requested by users, beds occupied with test-positive patients. This is generated by fitting a convolution between the admissions and occupancy time series, giving an estimate for time-to-discharge, translating the admissions forecast into an occupancy forecast.

To compare models and tune parameters we use probabilistic scoring methods, primarily the weighted interval score, empirical coverage, and bias, calculated using the *scoringutils* [33] R package. 90% and 50% prediction intervals are generated, with the 90% and median value are communicated to users, alongside probabilistic statements assessing trends. While quantitative scoring is a key component in defining and improving our ensemble, expert judgement in the modelling team is necessary to exclude models in a given week.

#### Benefits of the stacking ensemble approach

Variations on the unweighted average quantile approach to ensembling are widely used in epidemic forecasting [34] [25]. Instead, we preserve individual model sample predictions [35], then perform an unweighted quantile summary of draws across all models. This allows us to capture uncertainty in a granular way when summarising predictions and avoid relying on averaging or aggregating point estimates.

Though this prediction sample approach requires higher data volume than point estimates, as models are run within one team this has a limited impact on operations. By using a prediction sample stacking approach, we can produce granular forecasts at local levels then aggregate the sample draws to higher geographies, allowing for simple reconciliation across space, time, and other quantities of interest (such as age in the RSV model) [36]. It is important to our users that we produce national forecasts (for ministers), low geographies (for health protection teams), and their coherence is important for credibility.

Lastly, the use of prediction samples allows us to assign probabilities to different interpretable trend categories (stable, increase and decrease) using thresholds agreed with disease and operations experts, achievable at all geographies. We consider a change over two weeks of <20% to be stable, a positive change >20% as an increase and a negative change >20% as a decrease, similar to other experimental approaches worldwide [37].

## 3. Results

In a time of disrupted and uncertain seasonal patterns caused by the SARS-CoV-2 pandemic we chose to forecast our metrics over a forecast horizon window, showing how the metric is expected to develop. As a trade-off between long forecast horizons with high uncertainty and short horizons with lower utility, we chose a consistent 14-day horizon across all models. This provides enough foresight to be useful, with meaningfully small uncertainty, agreed with end users of the forecasts.

### 3.1 Season overview and targets

Each disease is monitored differently across the healthcare system, with different levels of granularity and quality available. The forecast targets selected for each disease and metric are given in **Table 1**. A key consideration in the choice of target data is its latency, traded off against its accuracy. For operational efficiency, forecasts for each disease were not delivered across every winter week. Forecasts were prioritised based on the trends observed, stopped early if needed, new models incorporated, and further metrics included as the season progressed. The timeline of metrics forecasted over the season are given in **Figure 1**.

**Figure 1.**
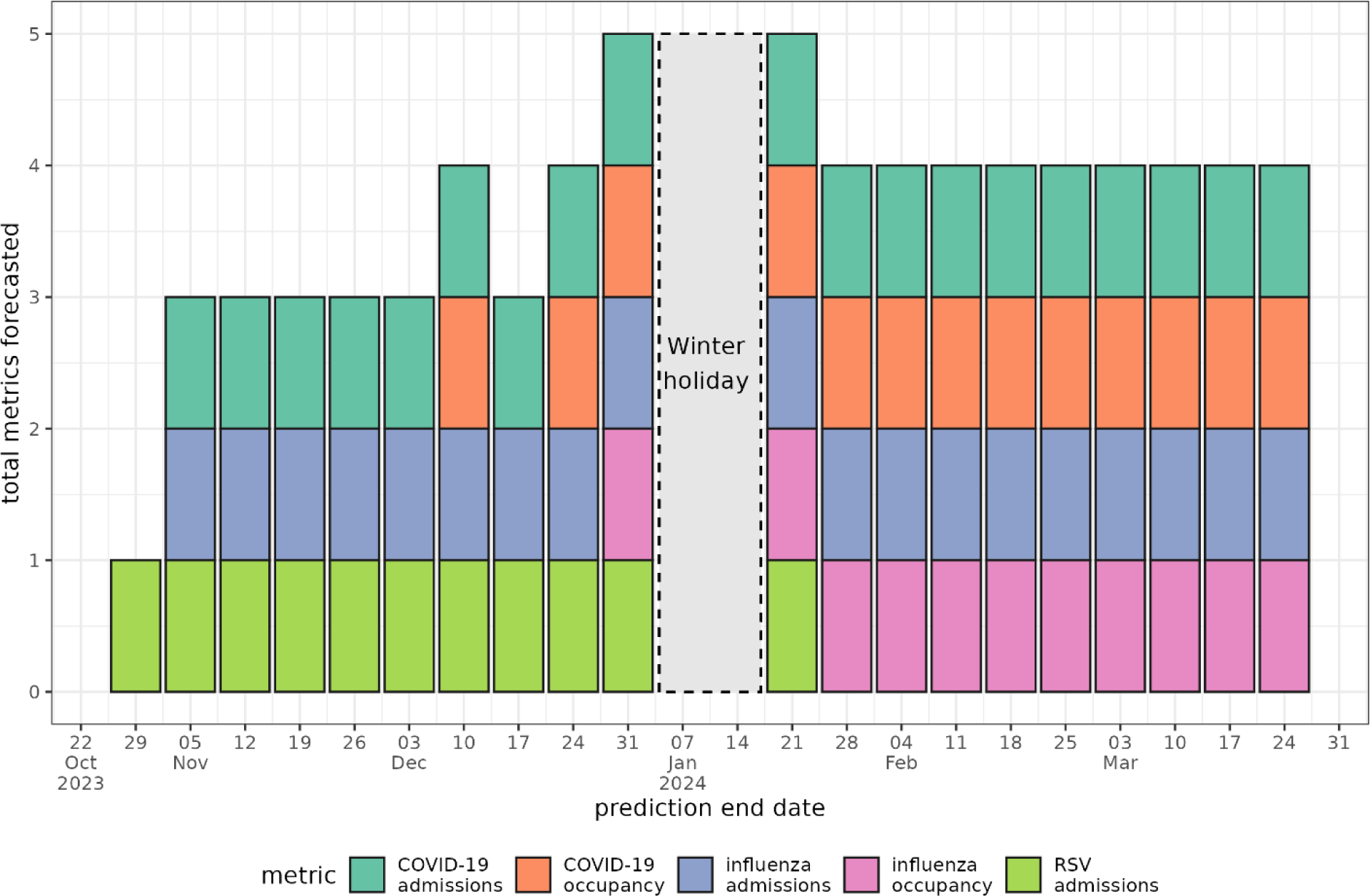
The weekly forecasts produced across pathogen and metric. There were no forecasts produced over the winter holiday period. Occupancy metrics began later in the season corresponding to higher pressure periods.

Our confidence in each data stream, and what is interesting to the users of our forecast varies by disease, and therefore the spatial resolution we model is not consistent across diseases. COVID-19 admissions have the highest quality of reporting, in part due to their high priority during the pandemic, though age information is no longer captured. Alternatively, for RSV, as individual data are processed by UKHSA, stratifications by age are possible allowing for models that incorporate demographic effects.

### 3.2 Influenza

The seasonal influenza epidemic in 2023/24 was characterized by a small peak before the New Year, and a second larger peak after (**Figure 2**). Observed 14-day admissions and occupancy values were within the forecasted 90% prediction intervals in 15/19 and 10/11 weeks respectively, though the admissions forecast performed poorly between peaks (**Figure 2**). For influenza, the GAM was included in the ensemble across all weeks, the ETS model incorporated later in the season, and leading indicator models used intermittently.

**Figure 2.**
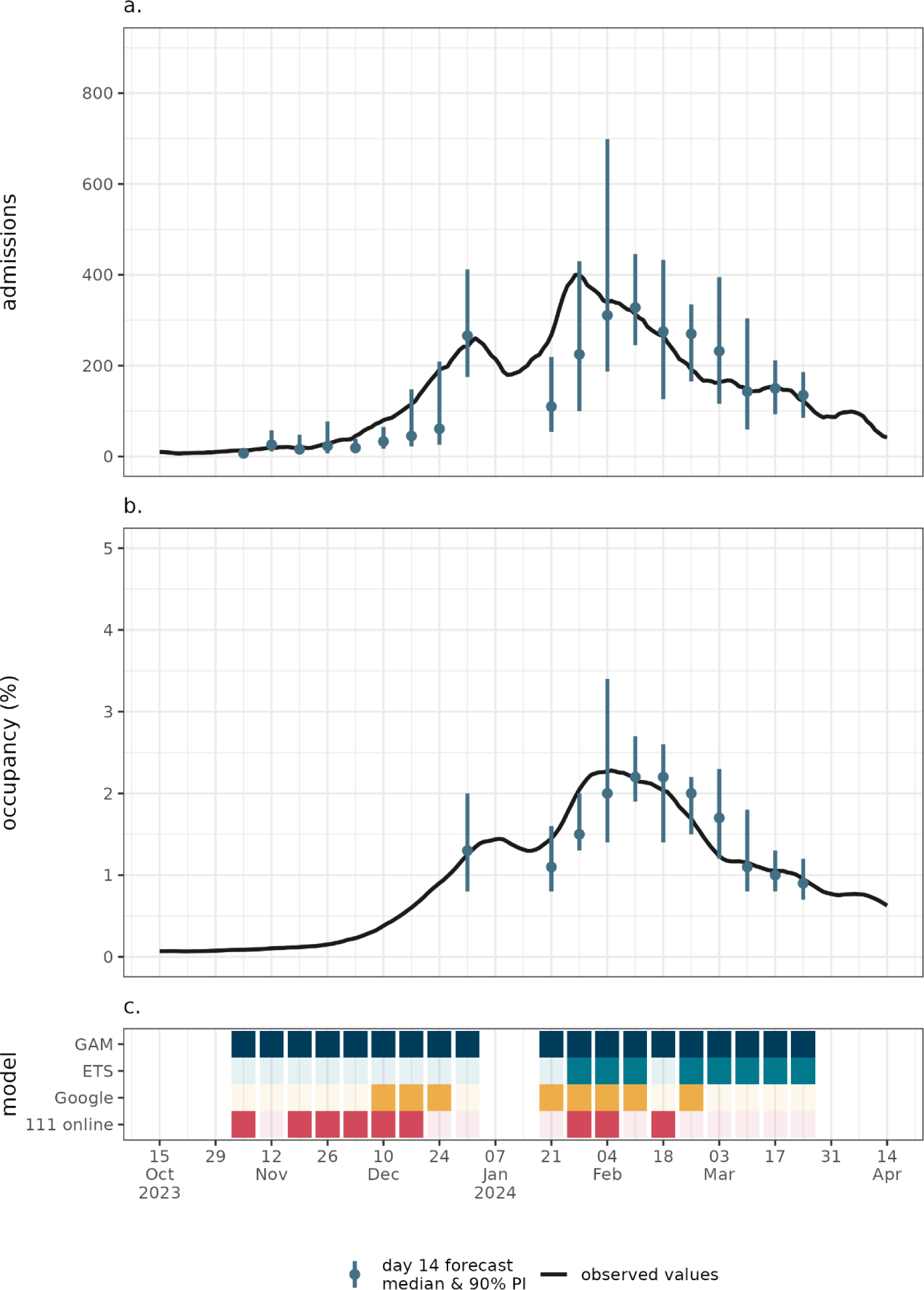
National level. a.) influenza admissions and b.) bed occupancy 14-day forecast overlaid. All metrics are 7-day rolling averaged for clarity. Plot c.) shows the models included in the ensemble each week.

### 3.3 COVID-19

Unlike influenza, there were COVID-19 admissions at non-zero levels at the start of winter, with multiple peaks and troughs observed in the admissions and occupancy trends (**Figure 3**). The main peak occurred near the New Year. The COVID-19 forecasts have variation in their accuracy, with forecasts generally struggling in growth/decline phases, where the models are slow to adapt to changing trends (**Figure 4**). Of the five available models for the ensemble, none were used across all weeks. The GAM was used most frequently, with sporadic use of leading indicator models, and the ETS model was regularly deployed towards the end of the season.

**Figure 3.**
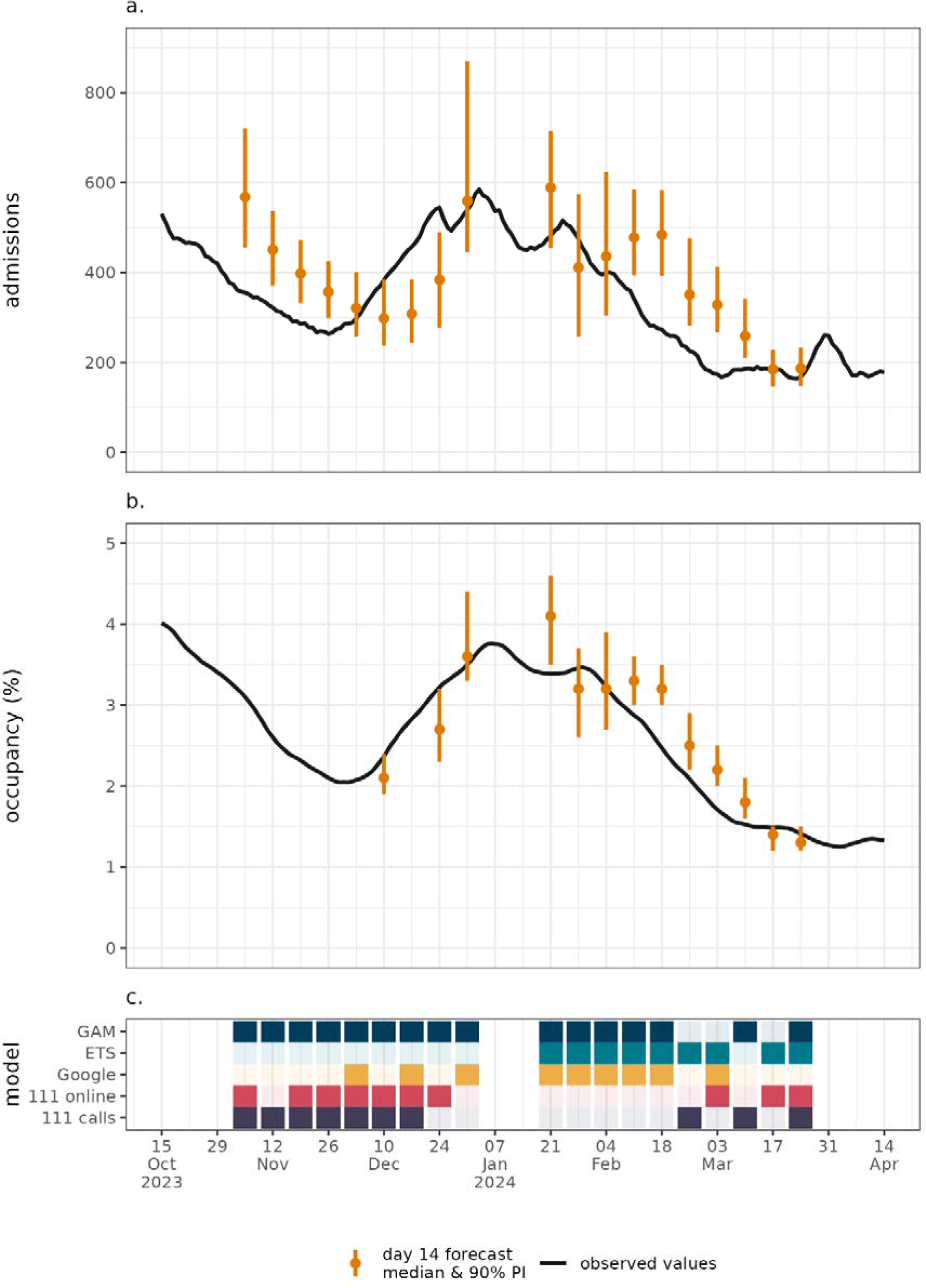
National level. a.) COVID-19 admissions and b.) bed occupancy 14 day forecast overlaid. All metrics are 7-day rolling averaged for clarity. Plot c.) shows the models included in the ensemble each week.

**Figure 4.**
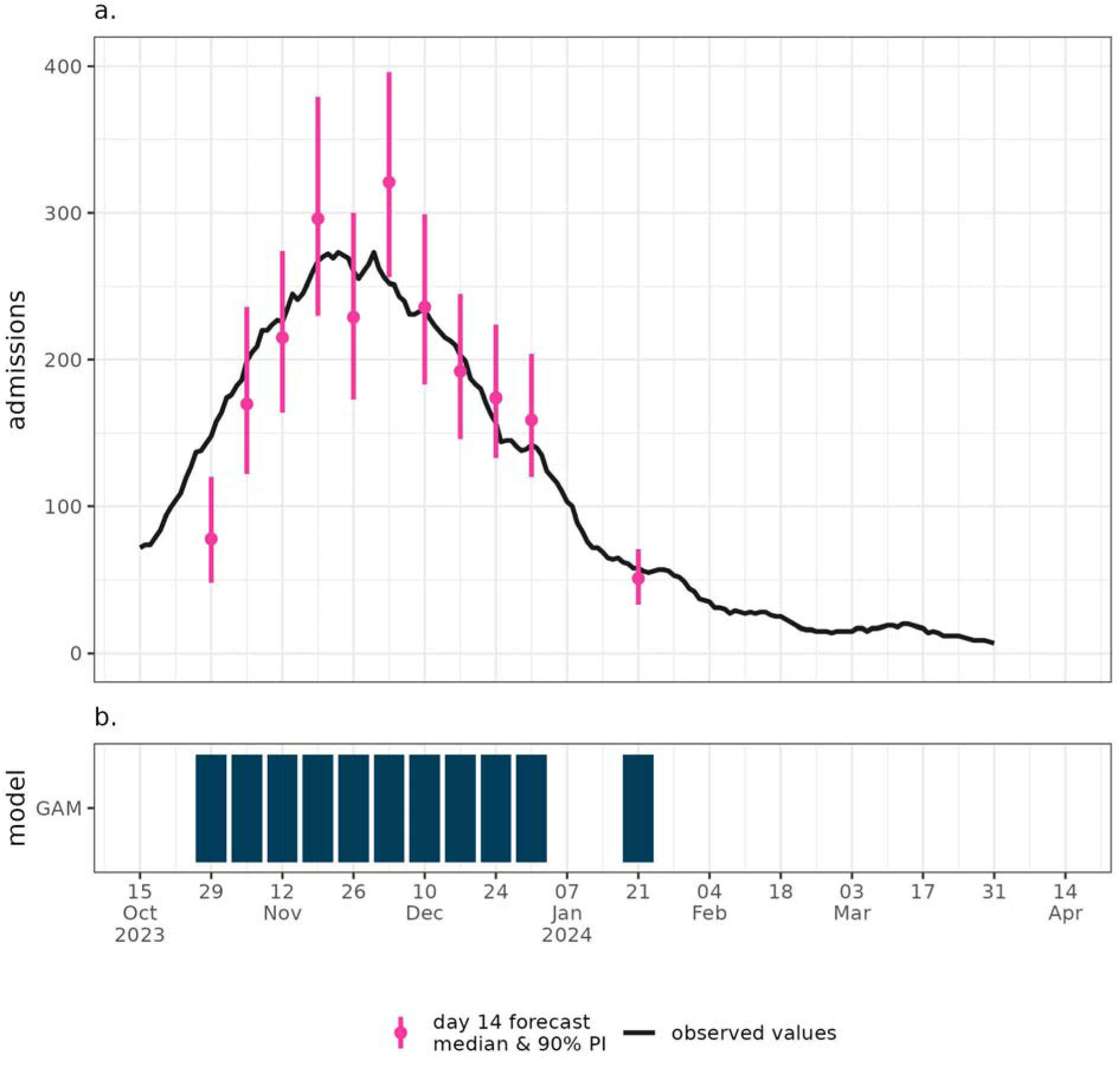
National level. a.) RSV admissions 14 day forecast overlaid. The metrics are 7-day rolling averaged for clarity. Plot b.) shows the models included each week. RSV forecasts were stopped early when incidence was low to prioritize other forecasts.

### 4.4 RSV

Relative to influenza and COVID-19, the peak in the RSV epidemic was earlier in the season, occurring in November (**Figure 4**). All but two 14-day forecasted 90% prediction intervals contain the observed values, capturing the trend in the epidemic well across the wave (**Figure 4**). One single model, the GAM with age and regional stratifications, was used to forecast this year.

### 4.5 Forecast performance

Quantitative evaluation of forecast accuracy is critical to the communication and improvement of forecasts. Measures of performance at a range of breakdowns show varying performance across diseases, metrics, and geographies (**Table 2**). Across 90% and 50% coverage scores the regional forecasts were better calibrated than their national aggregations. The occupancy forecasts for COVID-19 and influenza had higher performing central estimates than the corresponding admissions forecasts, a result of the slower evolving trend dependent on past admissions.

**Table 2.** Forecast scoring measures across disease, metric, and geographies. The 90% and 50% coverage metrics present the proportion of true values that fell within the specified prediction intervals, a measure of model calibration, with values closer to 0.9 or 0.5 preferred. For forecasts with true values of zero, a percentage error of 100% is assumed to define a consistent measure. The Weighted interval score accounts for model sharpness and bias, with lower scores being preferred. Each score is calculated over all dates predicted, for each weekly prediction.

| Disease | Metric | Geography | 90% coverage | 50% coverage | Percent of median estimates within 20% of true value | Mean absolute percentage error | Weighted interval score |
| --- | --- | --- | --- | --- | --- | --- | --- |
| COVID-19 | admissions | nation | 0.439 | 0.198 | 42.5 | 27.3 | 57 |
| COVID-19 | admissions | region | 0.835 | 0.422 | 40.7 | 34.0 | 8.2 |
| Influenza | admissions | nation | 0.807 | 0.438 | 39.1 | 30.9 | 26 |
| Influenza | admissions | region | 0.875 | 0.506 | 26.9 | 48.4 | 5.4 |
| RSV | admissions | nation | 0.779 | 0.397 | 72.5 | 15.7 | 16 |
| RSV | admissions | region | 0.859 | 0.514 | 43.2 | 37.7 | 3.9 |
| COVID-19 | occupancy | nation | 0.526 | 0.281 | 83.6 | 10.7 | 0.17 |
| COVID-19 | occupancy | region | 0.810 | 0.419 | 72.2 | 14.9 | 0.19 |
| Influenza | occupancy | nation | 0.889 | 0.556 | 80.2 | 10.0 | 0.087 |
| Influenza | occupancy | region | 0.861 | 0.517 | 65.8 | 18.9 | 0.15 |

The forecast performances can also be explored relative to the winter epidemic wave peak (**Figure 5**). For the influenza admissions, occupancy, and COVID-19 admissions there is a notable drop in coverage performance following the epidemic peak, indicating models may struggle to be calibrated at this point. Notably, as with the results in **Table 2**, the regional coverage is consistently better than at national levels, particularly for the COVID-19 models, even across epidemic phases. Further scoring metrics, such as the coverage deviation and bias are explored in **Supplementary Tables 2 & 3**.

**Figure 5.**
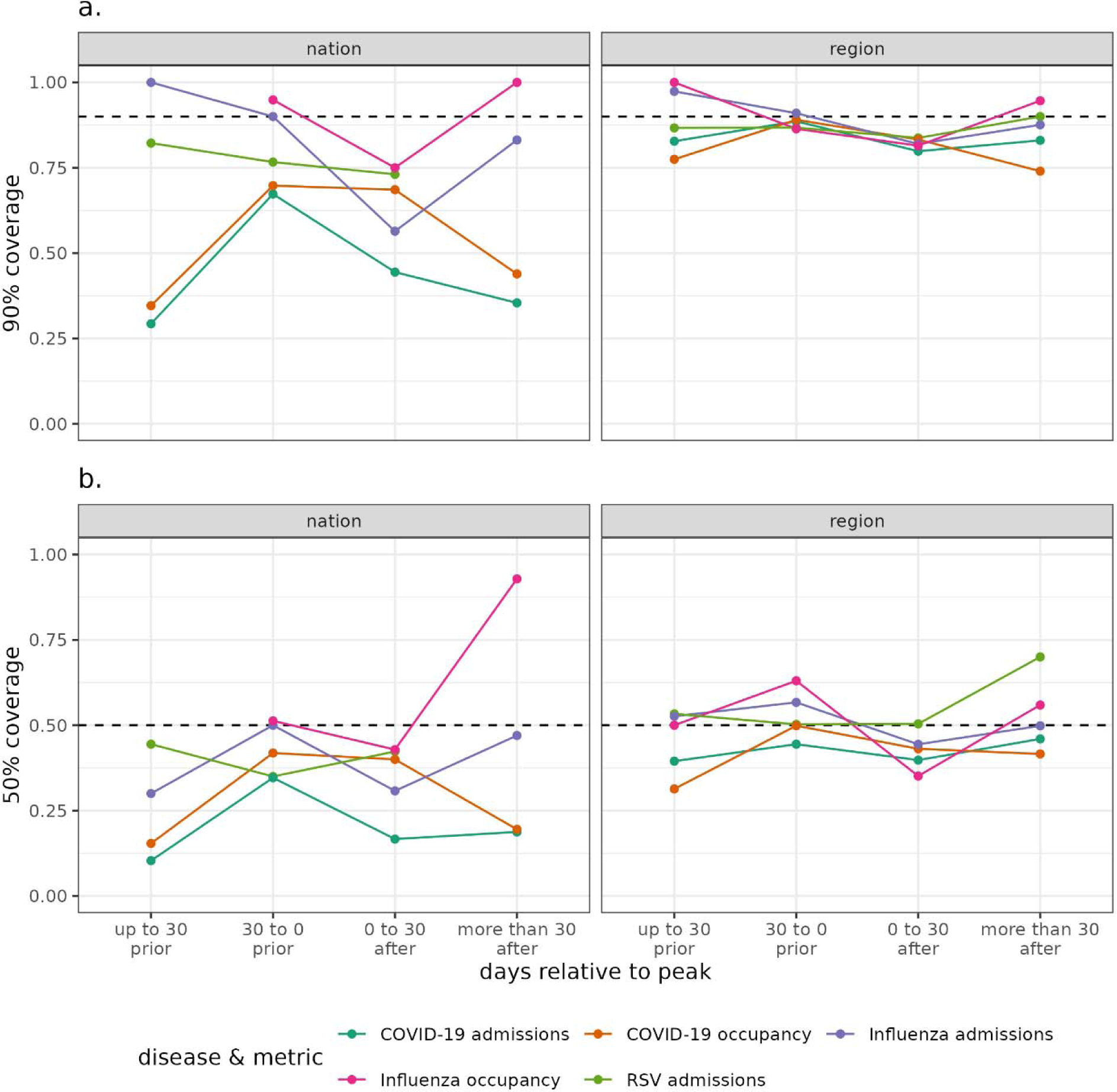
The. a.) 90% and b.) 50% empirical coverage of each disease and metric at national and regional geographies. The scores are given relative to the peak timing, to evaluate at different phases in the epidemic. Empirical coverage closer to the dashed line are preferred. The peak date is calculated as the date of maximum seven-day rolling average metric value over the winter period.

## 4. Discussion

In this manuscript we describe the approach taken by modellers at UKHSA to deliver a suite of forecasts across three priority diseases in the 2023/24 winter season. Forecasts harnessed the unified data collection of the health service in England, giving high coverage of hospitals, low latency, and granular spatio-temporal resolution. Some forecasts were particularly accurate over the season, such as the RSV admissions results (**Figure 4**). However, modelling for COVID-19 admissions were particularly challenging this season (**Figure 2**). The forecasts were widely disseminated within the health system, with modellers and operations colleagues collaborating closely. The resulting forecasts had high utility at both national and regional geographies, aiding decision makers in their resource allocation decisions.

Forecast users ranged from senior ministers responsible for the nation’s health, to regional health protection teams, with a variety of use-cases in between (**Figure 6**). National forecasts were shared each week with the Department of Health and Social Care to support oversight by the Secretary of State and ministers. This aided national-level decisions on system performance and operating policy over the winter, along with other analyses. At a sub-national level, forecasts were shared at weekly regional operational meetings held by NHS England. This gave foresight for regional decision-makers on hospital capacity, operational pressure [38], and coordination of mutual aid between hospitals and regions. Forecasts were also used by regional health protection teams to anticipate pressure on public health response.

**Figure 6.**
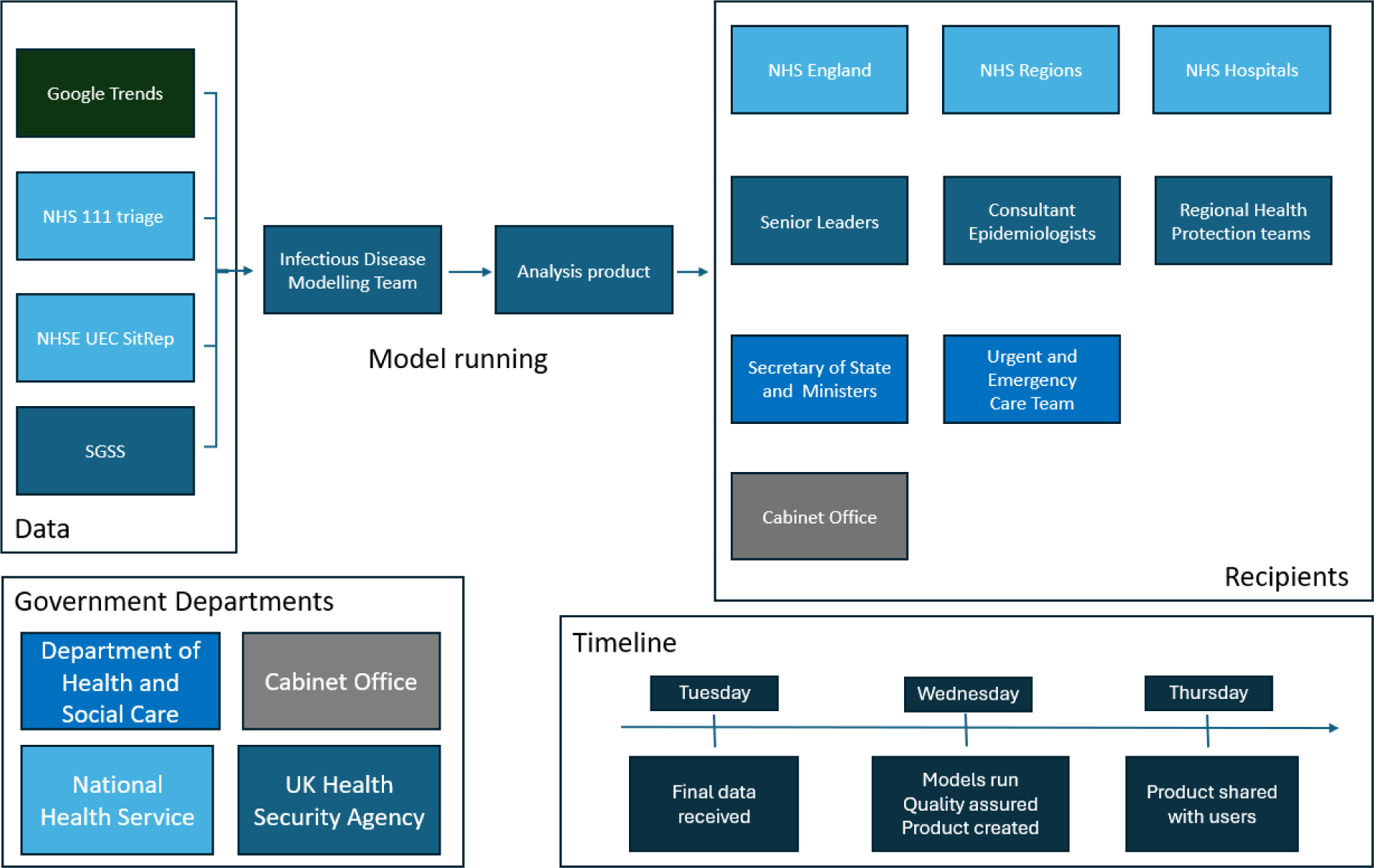
Flow chart of data sources and model dissemination across government decision makers in the 2023/24 season.

Developing a modelling suite in-house allowed UKHSA modellers to leverage existing relationships between disease experts and data collectors to develop useful forecasts. Granular, unified data collection enabled geography-specific forecasts to be created, directly supporting local operations, strengthening relationships between users and forecasters. By developing the forecasting suite within a single team, it was straightforward to create new targets and experiment with new approaches, which will be necessary in future outbreaks. By focusing on statistical time-series methods, the suite can be scaled to new diseases and metrics, without the challenging parameterisation of more mechanistic approaches. A range of model types can be used with the sample-based ensembling, giving strong reconciliation across geographic scales, and allowing for trend categorisation that aids decision making.

The modelling suite was developed iteratively over the past three years. It began in 2021 as a single COVID-19 model, then for winter 2022/23 evolved into a COVID-19 ensemble and a new influenza model. Most recently, in 2023/24 we created model ensembles for both COVID-19 and influenza, with an RSV model added. This build-up of models over time allowed adaption to evolving user needs, experimentation with new approaches, and streamlining processes in quieter periods.

The choice of forecasting target underpins the utility of the modelling, particularly the forecast horizon. Statistical models have been shown to excel at short horizons, relative to longer time periods [22]. Our work has a short-term focus, with a 14 day-look-ahead, providing more reasonable uncertainty in trends compared to other methods. Though, there is less room for decision makers to plan as the horizon shortens. We have found, while short, the 2-week horizon is highly useful for stakeholders, with meaningful uncertainty. Scenario modelling over longer term horizons has a different purpose to the immediate decision making of short-term forecasting, where academic collaborations have excelled [39].

The natural extensions to this modelling suite are clear: improvements in forecast accuracy, extending the forecast horizon, and exploration of other disease targets and further granularity. To further improve situational awareness, combinations of nowcasting and forecasting should be considered to harness inherently lagged data. The current suite of models is primarily statistical. Deploying models with more mechanistic approaches, including further disease dynamics assumptions, and incorporating historical trends are further avenues for exploration [40]. Further diseases should be considered for forecasting, creating the capability for responsive modelling work, though this necessitates improved timeliness of data collection, which currently exists only for influenza and COVID-19 in England. Increased sharing of data and code would benefit methods development and assurance, though we must be careful this does not limit modellers’ agility to deliver in response periods, and flexibility to try new targets and metrics.

The choice of modelling in-house only, largely due to data sharing challenges, limits the public availability of forecast information in real-time. The work being closed source, rather than code existing in the public domain, limits the scrutiny and collaboration available for the project. Furthermore, by focusing efforts within a single team there is a limit to time available for novel model development. This leaves expertise, primarily academic, both within England and internationally, that is not necessarily being leveraged to inform predictions – highlighting the importance of government and academic collaboration. The ensemble is limited by a lack of mechanistic dynamic components, such as susceptible population depletion, which may help with performance at epidemic turning points.

Over the 2023/24 winter season, we delivered a forecasting suite for respiratory diseases within secondary care in England with a range of statistical and ensemble models. These model outputs were used widely across the health system for situational awareness and supported decision making at national and local levels. We have shown that having an internal capability for real-time modelling of infectious diseases supports the delivery of effective public health.

## Acknowledgements

We would like to specifically thank members of the Infectious Disease Modelling team for their contributions to the operational delivery of the winter forecasts over the season:

– William Ferguson
– Jack Kennedy
– Emilie Finch
– Oliver Polhill
– Chetan Chauhan-Sims
– Adrian Pritchard
– Rachel Christie

In addition, we would like to thank the analysts within the NHS England Urgent and Emergency Care teams for their support, as well as colleagues within the NHS collecting and reporting the data used in this work.

## Ethical Approval

UKHSA have an exemption under regulation 3 of section 251 of the National Health Service Act (2006) to allow identifiable patient information to be processed to diagnose, control, prevent, or recognise trends in, communicable diseases and other risks to public health.

## Data Availability Statement

UKHSA⍰operates a robust governance process for applying to access protected data that considers:

- the benefits and risks of how the data will be used
- compliance with policy, regulatory and ethical obligations
- data minimisation
- how the confidentiality, integrity, and availability will be maintained
- retention, archival, and disposal requirements
- best practice for protecting data, including the application of “privacy by design and by default”, emerging privacy conserving technologies and contractual controls

Access to protected data is always strictly controlled using legally binding data sharing contracts.

UKHSA⍰welcomes data applications from organisations looking to use protected data for public health purposes.

To request an application pack or discuss a request for UKHSA data you would like to submit, contact.

## References

[1] UK Health Security Agency, “Surveillance of influenza and other seasonal respiratory viruses in the UK, winter 2023 to 2024,” 11 June 2024. [Online]. Available: https://www.gov.uk/government/statistics/surveillance-of-influenza-and-other-seasonal-respiratory-viruses-in-the-uk-winter-2023-to-2024/surveillance-of-influenza-and-other-seasonal-respiratory-viruses-in-the-uk-winter-2023-to-2024. [Accessed 5 September 2024].

[2] UK Health Security Agency, “National flu and COVID-19 surveillance reports: 2023 to 2024 season,” May 2024. [Online]. Available: https://www.gov.uk/government/statistics/national-flu-and-covid-19-surveillance-reports-2023-to-2024-season. [Accessed 5 September 2024].

[3] C. S. Lutz, M. P. Huynh, M. Schroeder, S. Anyatonwu, F. S. Dahlgren, G. Danyluk, D. Fernandez, S. K. Greene, N. Kipshidze and L. Liu, “Applying infectious disease forecasting to public health: a path forward using influenza forecasting examples,” BMC Public Health, vol. 19, pp. 1–12, 2019.

[4] C. J. McGowan, M. Biggerstaff, M. Johansson, K. M. Apfeldorf, M. Ben-Nun, L. Brooks, M. Convertino, M. Erraguntla, D. C. Farrow and J. Freeze, “Collaborative efforts to forecast seasonal influenza in the United States, 2015–2016,” Scientific reports, vol. 9, no. 1, p. 683, 2019.

[5] K. M. Holcomb, S. Mathis, J. E. Staples, M. Fischer, C. M. Barker, C. B. Beard, R. J. Nett, A. C. Keyel, M. Marcantonio and M. L. Childs, “Evaluation of an open forecasting challenge to assess skill of West Nile virus neuroinvasive disease prediction,” Parasites & Vectors, vol. 16, no. 1, p. 11, 2023.

[6] C. Viboud, K. Sun, R. Gaffey, M. Ajelli, L. Fumanelli, S. Merler, Q. Zhang, G. Chowell, L. Simonsen and A. Vespignani, “The RAPIDD ebola forecasting challenge: Synthesis and lessons learnt,” Epidemics, vol. 22, pp. 13–21, 2018.

[7] S. Y. Del Valle, B. H. McMahon, J. Asher, R. Hatchett, J. C. Lega, H. E. Brown, M. E. Leany, Y. Pantazis, D. J. Roberts and S. Moore, “Summary results of the 2014-2015 DARPA Chikungunya challenge,” BMC infectious diseases, vol. 18, pp. 1–14, 2018.

[8] S. M. Mathis, A. E. Webber, T. M. León, E. L. Murray, M. Sun, L. A. White, L. C. Brooks, A. Green, A. J. Hu and R. Rosenfeld, “evaluation of FluSight influenza forecasting in the 2021–22 and 2022–23 seasons with a new target laboratory-confirmed influenza hospitalizations,” Nature communications, vol. 15, 2024.

[9] N. G. Reich, C. J. McGowan, T. K. Yamana, A. Tushar, E. L. Ray, D. Osthus, S. Kandula, L. C. Brooks, W. Crawford-Crudell and G. C. Gibson, “Accuracy of real-time multi-model ensemble forecasts for seasonal influenza in the US,” PLoS computational biology, vol. 15, no. 11, p. e1007486, 2019.

[10] E. Y. Cramer, Y. Huang, Y. Wang, E. L. Ray, M. Cornell, J. Bracher, A. Brennen, A. J. C. Rivadeneira, A. Gerding and K. House, “The United States covid-19 forecast hub dataset,” Scientific data, vol. 9, no. 1, p. 462, 2022.

[11] E. Y. Cramer, E. L. Ray, V. K. Lopez, J. Bracher, A. Brennen, A. J. Castro Rivadeneira, A. Gerding, T. Gneiting, K. H. House and Y. Huang, “Evaluation of individual and ensemble probabilistic forecasts of COVID-19 mortality in the United States,” Proceedings of the National Academy of Sciences, vol. 119, no. 15, p. e2113561119, 2022.

[12] H. E. Groves, P.-P. Piché-Renaud, A. Peci, D. S. Farrar, S. Buckrell, C. Bancej, C. Sevenhuysen, A. Campigotto, J. B. Gubbay and S. K. Morris, “The impact of the COVID-19 pandemic on influenza, respiratory syncytial virus, and other seasonal respiratory virus circulation in Canada: A population-based study,” The Lancet Regional Health–Americas, vol. 1, 2021.

[13] S. S. Lee, C. Viboud and E. Petersen, “Understanding the rebound of influenza in the post COVID-19 pandemic period holds important clues for epidemiology and control,” International Journal of Infectious Diseases, vol. 122, pp. 1002–1004, 2022.

[14] A. J. Elliot, K. W. Cross and D. M. Fleming, “Acute respiratory infections and winter pressures on hospital admissions in England and Wales 1990–2005,” Journal of public health, vol. 30, no. 1, pp. 91–98, 2008.

[15] J. Mellor, R. Christie, J. Guilder, R. S. Paton, S. E. C. Watson, S. E. Deeny and T. Ward, “Influenza Hospitalisations in England during the 2022/23 Season: do different data sources drive divergence in modelled waves? A comparison of surveillance and administrative data,” 20 October 2023. [Online]. Available: https://www.medrxiv.org/content/10.1101/2023.10.19.23297248v1.

[16] S. Pei and J. Shaman, “Aggregating forecasts of multiple respiratory pathogens supports more accurate forecasting of influenza-like illness,” PLoS computational biology, vol. 16, no. 11, p. e1008301, 2020.

[17] Nation Health Service Digital, “Data collection and curation,” June 2024. [Online]. Available: https://digital.nhs.uk/data-and-information/data-collection-and-curation.

[18] C. Onwuchekwa, L. M. Moreo, S. Menon, B. Machado, D. Curcio, W. Kalina, J. E. Atwell, B. D. Gessner, M. Siapka and N. Agarwal, “Underascertainment of respiratory syncytial virus infection in adults due to diagnostic testing limitations: a systematic literature review and meta-analysis,” The Journal of infectious diseases, vol. 228, no. 2, pp. 173–184, 2023.

[19] E. Dietz, E. Pritchard, K. Pouwels, M. Ehsaan, J. Blake, C. Gaughan, E. Haduli, H. Boothe, K.-D. Vihta and T. Peto, “SARS-CoV-2, influenza A/B and respiratory syncytial virus positivity and association with influenza-like illness and self-reported symptoms, over the 2022/23 winter season in the UK: a longitudinal surveillance cohort,” BMC medicine, vol. 22, no. 1, p. 143, 2024.

[20] S. K. Sahu, B. Baffour, P. R. Harper, J. H. Minty and C. Sarran, “A hierarchical Bayesian model for improving short-term forecasting of hospital demand by including meteorological information,” Journal of the Royal Statistical Society Series A: Statistics in Society, vol. 177, no. 1, pp. 39–61, 2014.

[21] N. G. Reich, L. C. Brooks, S. J. Fox, S. Kandula, C. J. McGowan, E. Moore, D. Osthus, E. L. Ray, A. Tushar and T. K. Yamana, “A collaborative multiyear, multimodel assessment of seasonal influenza forecasting in the United States,” Proceedings of the National Academy of Sciences, vol. 116, no. 8, pp. 3146–3154, 2019.

[22] C. Viboud and A. Vespignani, “The future of influenza forecasts,” Proceedings of the National Academy of Sciences, vol. 116, no. 8, pp. 2802–2804, 2019.

[23] J. K. Sivillo, J. E. Ahlquist and Z. Toth, “An ensemble forecasting primer,” Weather and forecasting, vol. 12, no. 4, pp. 809–818, 1997.

[24] S. Meakin, S. Abbott, N. Bosse, J. Munday, H. Gruson, J. Hellewell, K. Sherratt and S. Funk, “Comparative assessment of methods for short-term forecasts of COVID-19 hospital admissions in England at the local level,” BMC Medicine, vol. 20, no. 1, p. 86, 2022.

[25] K. Sherratt, H. Gruson, H. Johnson, R. Niehus, B. Prasse, F. Sandmann, J. Deuschel, D. Wolffram, S. Abbott and A. Ullrich, “Predictive performance of multi-model ensemble forecasts of COVID-19 across European nations,” eLife, vol. 12, p. e81916, 2023.

[26] Y. Yao, A. Vehtari, D. Simpson and A. Gelman, “Using stacking to average Bayesian predictive distributions (with discussion),” Bayesian Analysis, vol. 13, no. 3, pp. 917–1003, 2018.

[27] J. Mellor, R. Christie, C. E. Overton, R. S. Paton, R. Leslie, M. Tang, S. Deeny and T. Ward, “Forecasting influenza hospital admissions within English sub-regions using hierarchical generalised additive models,” Nature Communications Medicine, vol. 3, no. 1, p. 190, 2023.

[28] S. N. Wood, “Fast stable restricted maximum likelihood and marginal likelihood estimation of semiparametric generalized linear models,” Journal of the Royal Statistical Society Series B: Statistical Methodology, vol. 73, no. 1, pp. 3–26, 2011.

[29] G. L. Simpson, “gratia: Graceful ggplot-Based Graphics and Other Functions,” July 2024. [Online]. Available: https://cran.r-project.org/web/packages/gratia/index.html.

[30] S. Meakin and S. Funk, “Quantifying the impact of hospital catchment area definitions on hospital admissions forecasts: COVID-19 in England, September 2020–April 2021,” BMC medicine, vol. 22, no. 1, p. 163, 2024.

[31] M. O’Hara-Wild, R. Hyndman and E. Wang, “fable: Forecasting Models for Tidy Time Series,” March 2024. [Online]. Available: https://cran.r-project.org/web/packages/fable/index.html.

[32] J. Mellor, C. E. Overton, M. Fyles, L. Chawner, J. Baxter, T. Baird and T. Ward, “Understanding the leading indicators of hospital admissions from COVID-19 across successive waves in the UK,” Epidemiology & Infection, vol. 151, p. e172, 2023.

[33] N. Bosse, S. Abbott, H. Gruson, J. Bracher and S. Funk, “scoringutils: Utilities for Scoring and Assessing Predictions,” 29 November 2023. [Online]. Available: https://cran.r-project.org/web/packages/scoringutils/index.html.

[34] J. W. Taylor and K. S. Taylor, “Combining probabilistic forecasts of COVID-19 mortality in the United States,” European Journal of Operational Research, vol. 304, no. 1, pp. 25–41, 2023.

[35] K. Sherratt, A. Srivastava, K. Ainslie, D. E. Singh, A. Cublier, M. C. Marinescu, J. Carretero, A. C. Garcia, N. Franco and L. Willem, “Characterising information gains and losses when collecting multiple epidemic model outputs,” Epidemics, vol. 47, p. 100765, 2024.

[36] R. Hollyman, F. Petropoulos and M. E. Tipping, “Understanding forecast reconciliation,” European Journal of Operational Research, vol. 294, no. 1, pp. 149–160, 2021.

[37] CDC Epidemic Prediction Initiative, “FluSight Forecast Data,” 2022. [Online]. Available: https://github.com/cdcepi/Flusight-forecast-data/tree/master/data-experimental. [Accessed July 2023].

[38] NHS England, “Operational Pressures Escalation Levels (OPEL) Framework 2023/24,” October 2023. [Online]. Available: https://www.england.nhs.uk/publication/operational-pressures-escalation-levels-opel-framework-2023-24/. [Accessed 28 August 2024].

[39] E. Howerton, L. Contamin, L. C. Mullany, M. Qin, N. G. Reich, S. Bents, R. K. Borchering, S.-m. Jung, S. L. Loo and C. P. Smith, “Evaluation of the US COVID-19 Scenario Modeling Hub for informing pandemic response under uncertainty,” Nature Communications, vol. 14, no. 1, p. 7260, 2023.

[40] S. Kandula, T. Yamana, S. Pei, W. Yang, H. Morita and J. Shaman, “Evaluation of mechanistic and statistical methods in forecasting influenza-like illness,” Journal of The Royal Society Interface, vol. 15, no. 144, p. 20180174, 2018.

