## Supplementary Information for "Forecasting COVID-19, Influenza and RSV hospitalisations over winter 2023/24 in England"

**Supplementary Table 1**. A breakdown of which models were used over the winter 2023/24 season. Diseases where there has been a longer standing forecast offering have larger ensembles developed, as the suite is iteratively improved. This season the 111 Calls telephonic health triage data source was only available for COVID-19 related triages.

| **Model** | **COVID-19** | **Influenza** | **RSV** |
| --- | --- | --- | --- |
| GAM | *Yes* | *Yes* | *Yes* |
| ETS | *Yes* | *Yes* | No |
| Regression: Google Trends | *Yes* | *Yes* | No |
| Regression: 111 Online | *Yes* | *Yes* | No |
| Regression: 111 Calls | *Yes* | No | No |

**Supplementary Table 2**. Bias related forecast scoring measures across disease, metric, and geographies.

| **Disease** | **Metric** | **Geography** | **Bias** | **Underprediction** | **Overprediction** |
| --- | --- | --- | --- | --- | --- |
| COVID-19 | admissions | nation | 0.31 | 12 | 33 |
| COVID-19 | admissions | region | 0.17 | 1.8 | 2.8 |
| Influenza | admissions | nation | -0.25 | 12 | 4.1 |
| Influenza | admissions | region | -0.21 | 2.5 | 0.63 |
| RSV | admissions | nation | 0.12 | 5.0 | 5.2 |
| RSV | admissions | region | 0.034 | 1.2 | 1.2 |
| COVID-19 | occupancy | nation | 0.36 | 0.017 | 0.12 |
| COVID-19 | occupancy | region | 0.22 | 0.028 | 0.078 |
| Influenza | occupancy | nation | 0.029 | 0.033 | 0.012 |
| Influenza | occupancy | region | -0.093 | 0.059 | 0.03 |

**Supplementary Table 3**. Forecast scoring rules across disease, metric, and geographies.

| **Disease** | **Metric** | **Geography** | **Weighted interval score** | **Dispersion** | **Coverage deviation** | **Median absolute error** |
| --- | --- | --- | --- | --- | --- | --- |
| COVID-19 | admissions | nation | 57 | 12 | -0.35 | 92 |
| COVID-19 | admissions | region | 8.2 | 3.6 | -0.064 | 15 |
| Influenza | admissions | nation | 26 | 9.7 | -0.062 | 52 |
| Influenza | admissions | region | 5.4 | 2.2 | -0.013 | 9.8 |
| RSV | admissions | nation | 16 | 5.5 | -0.085 | 29 |
| RSV | admissions | region | 3.9 | 1.7 | -0.0013 | 7.3 |
| COVID-19 | occupancy | nation | 0.17 | 0.039 | -0.27 | 0.28 |
| COVID-19 | occupancy | region | 0.19 | 0.083 | -0.074 | 0.36 |
| Influenza | occupancy | nation | 0.087 | 0.042 | 0.014 | 0.17 |
| Influenza | occupancy | region | 0.15 | 0.065 | -0.012 | 0.27 |

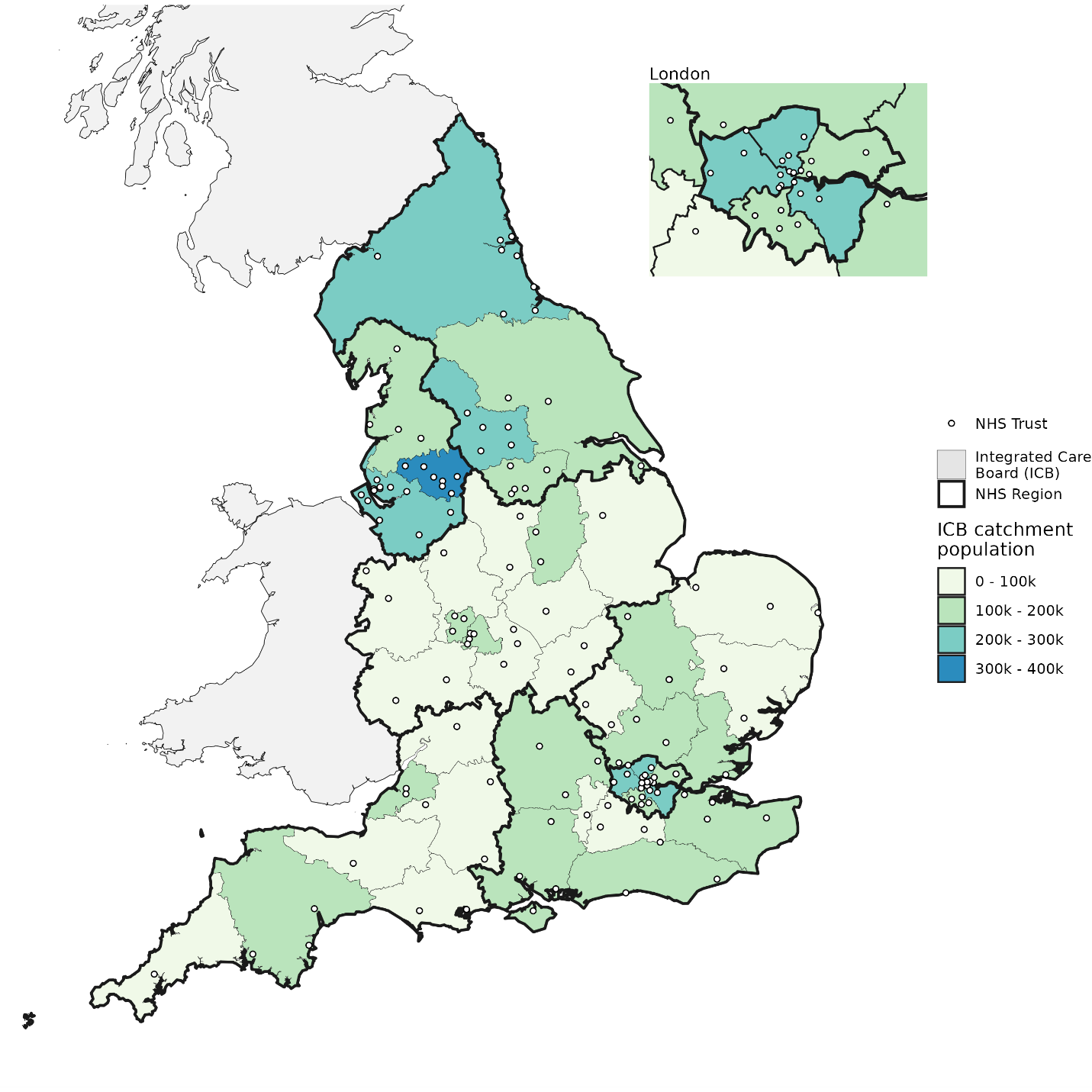
**Supplementary Figure 1.** Map of England with NHS geographies shown. Each acute NHS Trust is shown as a point location based on its headquarters postcode. Each Integrated Care Board (ICB) has a fixed boundary, with one or multiple NHS Trusts within its border, though they treat patients across borders. The 7 NHS regions contain multiple ICBs each.
